# Control Strategies to Curtail Transmission of COVID-19

**DOI:** 10.1101/2020.04.04.20053173

**Authors:** Nita H. Shah, Ankush H. Suthar, Ekta N. Jayswal

## Abstract

Recently the World Health Organization has declared the outbreak of a severe acute respiratory syndrome coronavirus 2 (SARS-Cov-2) as a pandemic, and declared it as Public Health Emergency of International Concern. More than 683,536 positive cases and 32,139 deaths caused by novel corona virus 2019 (COVID-19) has affected 199 countries and territories. This pandemic can transform into an extremely destructive form if we still do not take it seriously. In the present study, we propose a generalized SEIR model of COVID-19 to study the behaviour of its transmission under different control strategies. In the model, all possible cases of human to human transmission are taken care and its reproduction number is formulated to analyse accurate transmission dynamics of the coronavirus outbreak. Optimal control theory is applied in the model to pretend the impact of various intervention strategies, including voluntary quarantine, isolation of infected individuals, improving an individual’s immunity and hospitalisation. Also, effect of the control strategies on model is analysed graphically by simulating the model numerically.

## 1. Introduction

In December, 2019, the pandemic outbreak of a novel coronavirus disease named COVID-19 raised intense attention not only within China but internationally [11]. Doctors and scientists tested the previously developed drugs to treat the infected people, but that failed to succeed. Then, in February, 2020, the World Health Organization (WHO) declared the outbreak of this highly contagious COVID-19 as pandemic globally [13]. To control the human to human virus transmission, the central government of China as well as all local governments had tightened preventive measures. However, the virus had spread rapidly across most of the regions in China and in other countries and territories around the world. One major cause of the quick spread of COVID-19 is the lack of information and awareness about the virus during its early stages of infection. As on March 29, 2020, 13:29 GMT, has affected 199 countries and territories around the world with 683,536 confirmed cases, of which 32,139 have passed away, and 25,422 are serious or critical (worldometers.info). Still there is a possibility that spread of this virus could be more intense and cause high mortality. While the New Year was enjoying their spring vacations, the outbreak of COVID-19 has accelerated, as most of the people were on their way to hometown or else travelling to different places for relaxation.

Symptoms of COVID-19 takes at least 2 to 10 days, which makes it is tough to isolate infected individuals during initial stage of the infection. The major symptoms of COVID-19 include dry coughing, high fever with difficulties in breathing [16]. The virus may spread in the environment through respiratory droplets of infected individuals when they cough or sneeze. Further an unaffected population becomes infected when they are exposed by touching the infected surface or while breathing in an infected environment [10]. During the initial stages of COVID-19 outbreak, such human transmissions were taking place because, wide-range of public was unaware of these risk factors, and the infected individuals were also not isolated and were spreading the virus unknowingly to other individuals. Moreover, the risk factor of contamination is very high since the virus can remain viable in environment for several days in favourable conditions [3, 12]. Several studies reveal that old-aged people, children and those with major diseases are having low immunity and tend to seriously affected once they become infected [8, 17]. Still, we lack with any proper treatment or vaccines as a cure for this disease. Hence to control its transmission further, isolating the infected individuals in special quarantine cells has been implemented in most of the countries. Despite of these prevention strategies, we are in danger as the transmission is still ongoing and the mortality due to the virus maintains a high level.

To combat this situation, studies like mathematical modelling play a crucial role to understand the pandemic behaviour of the infectious disease. Several studies have already been undertaken to analyse the COVID-19 transmission dynamics. Based on these database studies of COVID-19 outbreak since 31^st^ December, 2019 to 28^th^ January, 2020, Wu *et. al*. (2020) introduced a SEIR-model to estimate the spread of the disease nationally as well as globally [18]. Tang *et. al*. (2020) proposed a compartmental model by dividing each group into two subpopulations, the quarantined and unquarantined. Moreover, they re-designed their previous model by using diagnose and time-dependent contact rates, and re-estimated the reproduction ratio to quantify the evolution in better way [14, 15]. Peng *et. al*. (2020) planned a generalized SEIR model that suitably incorporates the intrinsic impact of hidden exposed and infectious cases of COVID-19 [9]. Chen *et. al*. (2020) presented the incubation period behaviour of local outbreak of COVID-19 by constructing a dynamic system [2]. Khan & Atangana (2020) described brief details of interaction among the bats, unknown hosts, humans and the infections reservoir by formulating the mathematical results of the mathematical fractional model [7]. Chen *et. al*. (2020) have also developed a Bats-Hosts-Reservoir-People transmission network model for simulating the potential transmission of COVID-19 [1]. Zhao *et. al*. (2020) divided susceptible people from Wuhan City into different age groups and developed a SEIARW-model based on market to person and person to person transmission routes [20]. Zhong *et. al*. (2020) constructed a mathematical model using epidemiological data and examined characteristics of the historical epidemic to make an early prediction of the 2019-nCoV outbreak in China [21]. Yang & Wang (2020) have studied the multiple transmission pathways in the infection dynamics and formed a mathematical model that describes the role of the environmental reservoir in the transmission and spread of COVID-19 [19].

In this work, a COVID-19 model is constructed to study human to human transmission of the virus in section 2. Optimal control theory is introduced and applied to the model to develop in section 3 and in section 4, the model is simulated numerically to observe effect of control strategies on the model.

## 2. COVID-19 Model Formulation

To analyse the human to human transmission dynamics of COVID-2019, a compartmental model is constructed. The model consists all possible human to human transmission of the virus. The COVID-2019 is highly contagious in nature and infected cases are seen in most of the countries around the world, hence in the model the susceptible population class is ignored and whole population is divided in five compartments, class of exposed individuals *E*(t) (individuals surrounded by infection by not yet infected), class of infected individuals by COVID-19 I(t), class of critically infected individuals by COVID-19 *C*(t), class of hospitalised individuals *H*(t) and class of dead individuals due to COVID-19 *D*(t). Human to human transmission dynamics of COVID-19 is describe graphically in figure 1. Parameters used in the model are described in table 1.

**Figure 1.**
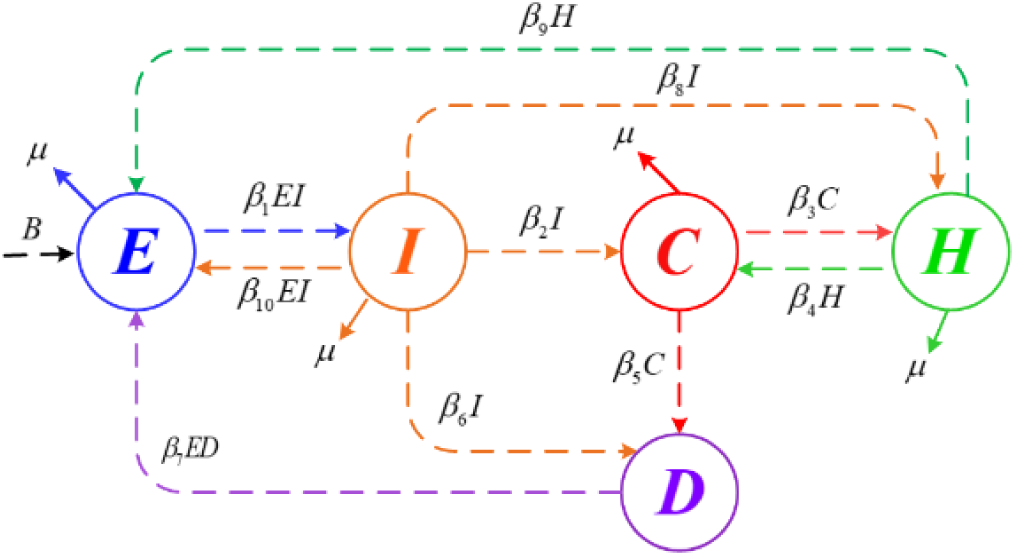
Transmission dynamics of COVID-19

**Table 1.** Parameters used in the model

| *Parameters* |  |  |  |
| --- | --- | --- | --- |
| $B$ | Birth rate of class of exposed individuals | 0.80 | Calculated |
| $\mu$ | Natural death rate | 0.01 | Assumed |
| $\beta_1$ | Transmission rate of individuals moving from exposed to infected class | 0.55 | Calculated |
| $\beta_2$ | Rate at which infected individuals goes into sever condition or in critical condition | 0.40 | Calculated |
| $\beta_3$ | Rate at which critically infected individuals get hospitalized | 0.60 | Calculated |
| $\beta_4$ | Rate by which hospitalized individuals not recovered and remain in critical condition | 0.80 | Calculated |
| $\beta_5$ | Mortality rate of critically infected individuals | 0.34 | Calculated |
| $\beta_6$ | Mortality rate of infected individuals | 0.30 | Calculated |
| $\beta_7$ | Rate by which infected dead body spreads infection | 0.35 | Assumed |
| $\beta_8$ | Rate at which infected individuals get hospitalized | 0.30 | Calculated |
| $\beta_9$ | Rate at which hospitalised individuals get recovered and become exposed again | 0.35 | Assumed |
| $\beta_{10}$ | Rate at which infected individuals recovered themselves due to strong immunity and again become exposed | 0.32 | Assumed |

Using the above depiction, dynamical system of set of nonlinear differential for the model is formulated as follows:

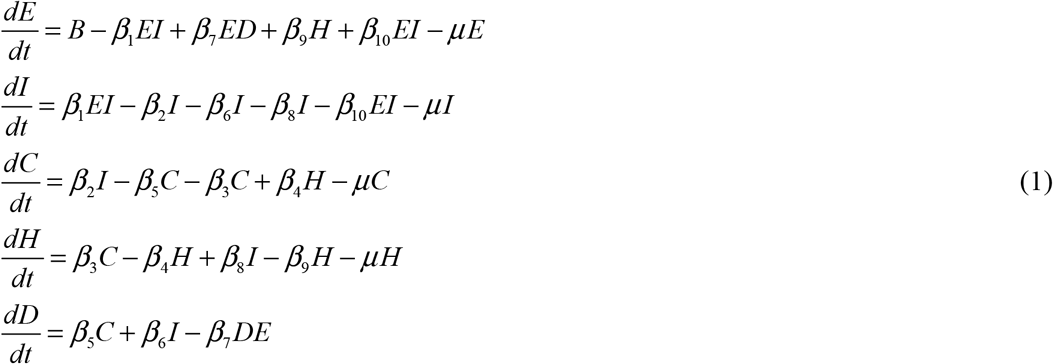

Note that, all the parameters used in this COVID-19 model are non-negative. Consider the feasible region,

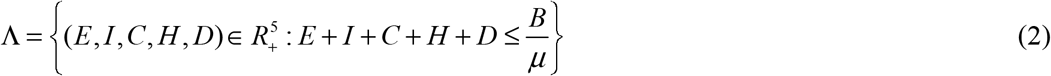

The region Λ is positively invariant, all the solutions of the system (1) are remain in the feasible region (2).

### 2.1. Equilibrium points

By solving the above system (1), we get two equilibrium points:

I. Disease free equilibrium point: 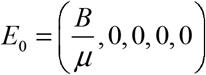
II. Endemic equilibrium point: 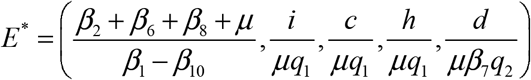

where,

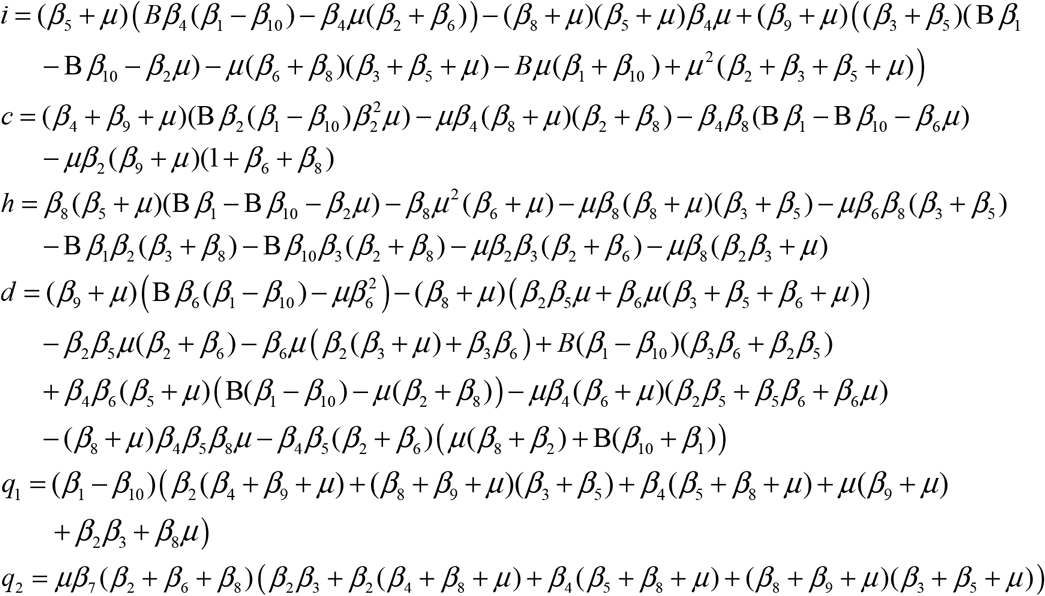

### 2.2. Basic reproduction number

Basic reproduction number (*R*_0_) for the model can be established using the next generation matrix method [4, 5]. The basic reproduction number (*R*_0_) is obtained as the spectral radius of matrix (*FV* ^−1^) at disease free equilibrium point. Where *F* and *V* are as below:

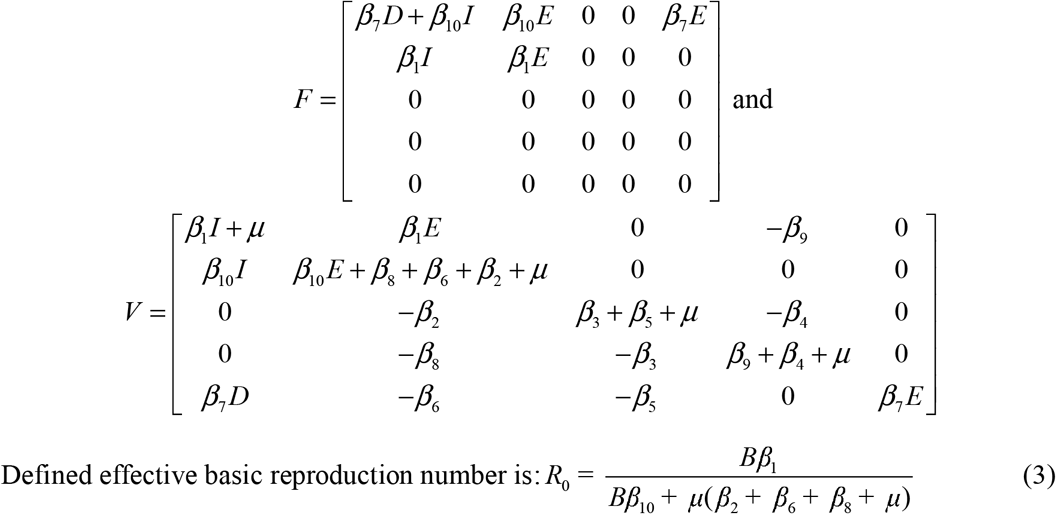

## 3. Optimal Control Theory

Control measures have made significant role to control the epidemic of COVID-19 at certain level. In this control theory, five control variables are used as five possible control strategies. Since the virus is highly contagious, it quickly infect any people come in contact with an infected individual. To avoid this situation we have taken *u*_1_ control variable to self-quarantine exposed individuals and *u*_2_ control variable as an isolation of infected individuals. Moreover, to minimise mortality rate of COVID-19, *u*_3_ control variable is taken which helps to reduce critically infected cases by taking extra medical care of infected individuals. *u*_4_ and *u*_5_ control variables are taken to improve hospitalisation facility for infected and critically infected individuals respectively. Purpose of this study of control theory is to protect people from the outbreak by applying control or treatment in each stage. Objective function for the required scenario is,

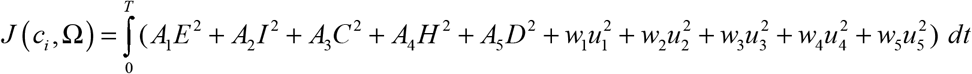

where, Ω denotes the feasible region for the set of compartmental variables, *A*_1_, *A*_2_, *A*_3_, *A*_4_, *A*_5_ denote non-negative weight constants for compartments *E, I,C, H, D* respectively. *w*_1_, *w*_2_, *w*_3_, *w*_4_ and *w*_5_ are the weight constants for each control *u*_*i*_, where *i* =1, 2,…,5 respectively. The modified model is,

Hence modified system for Fig. (2) is stated as:

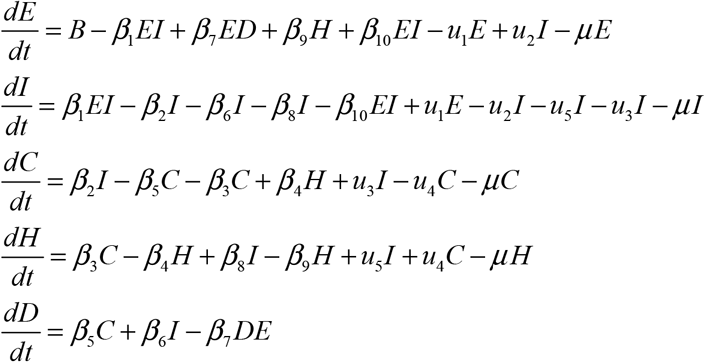

As the weight parameter *w*_*i*_, *i* =1, 2,…,5 are constants applied on the control variable respectively from which the optimal condition is normalized. Now, calculate every values of control variables from *t* = 0 to *t* = *T* such that,

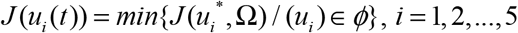

where, *ϕ* is a smooth function on the interval [0,1]. The optimal effect is found by using results of Fleming and Rishel (2012) [6]. Associated Langrangian function with adjoint variable *λ*_1_, *λ*_2_, *λ*_3_, *λ*_4_, *λ*_5_ is given by,

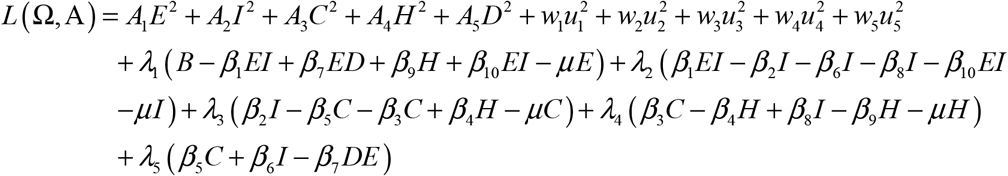

The partial derivatives of the Lagrangian function with respect to each variable of the compartment gives the adjoint equation variables system which are: *A*_*i*_ = (*λ*_1_, *λ*_2_, *λ*_3_, *λ*_4_, *λ*_5_) corresponding to the system which are:

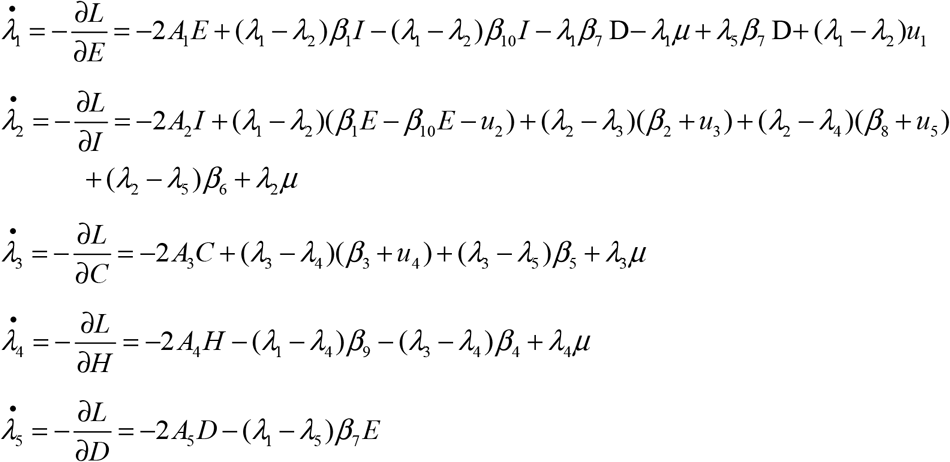

Hence, the optimal controls are given by

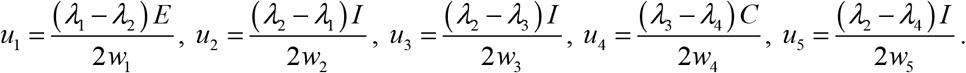

And optimal conditions given as,

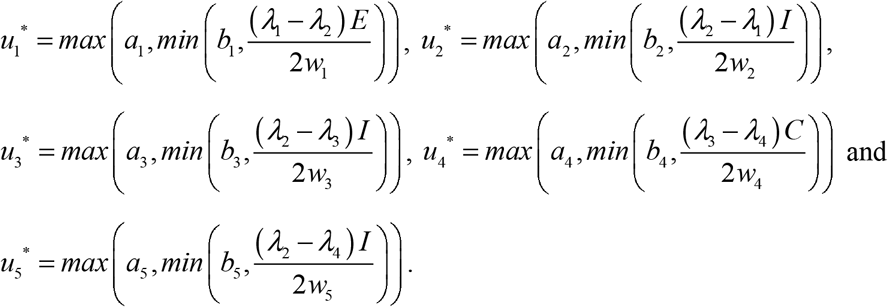

This calculation gives analytical behaviour of optimal control on the system. Numerical interpretation of optimal control theory is simulated in the next section.

## 4. Numerical Simulation

[1] In this section, the COVID-19 model is simulated numerically, wherein the parametric values for simulation are taken from recent pandemic outbreak of coronavirus (https://www.who.int/emergencies/diseases/novel-coronavirus-2019/situation-reports).

Figure 3 represents the variations in all the compartments of COVID-19 model with respect to time. Also, the pandemic behaviour of the COVID-19 outbreak can be clearly seen here. We can say that, a large population of exposed individuals become infected before a week. Moreover, the critically infected cases and hospitalisation cases also increase with matter of time. Further, it clearly shows that after one week the mortality rate is also increased.

**Figure 2.**
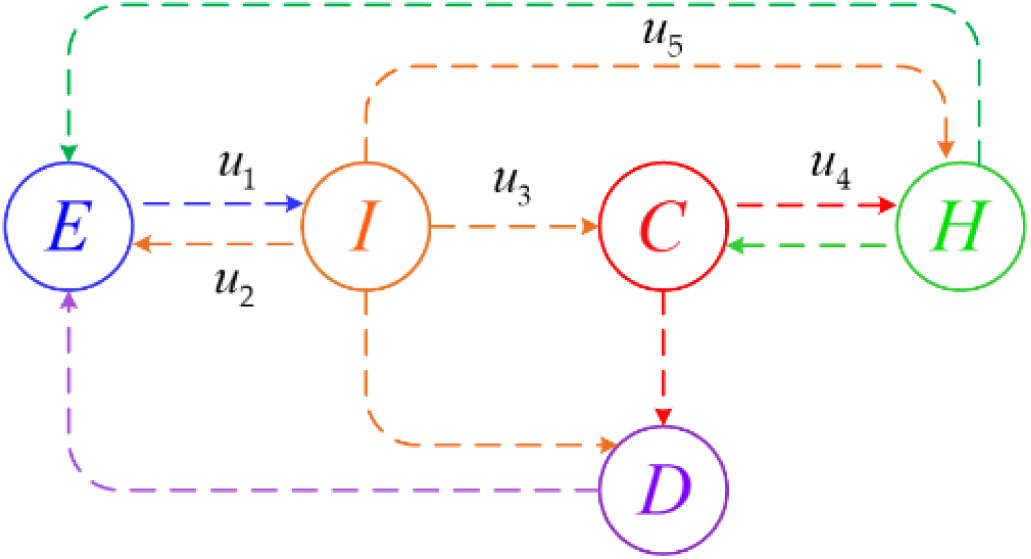
COVID-19 model with control variables

**Figure 3.**
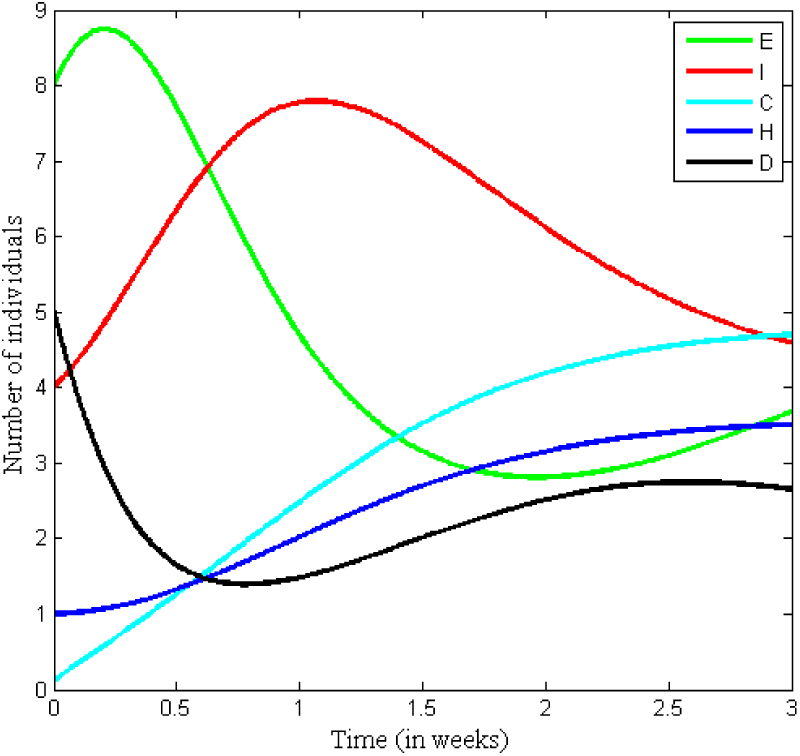
variation in all compartments

Figure 4 (a) shows variation in each compartment under influence with and without control variables. It is observed that COVID-19 outbreak can be controlled up to a certain level in seven weeks after applying control strategies. While, figures 4 (b) and (c) represent number of infected and critically infected individuals decreasing; which is further leading to a decrease in hospitalised class as shown in figure 4 (d). Thus, reduction in mortality rate under the effect of control strategies can be clearly observed in figure 4 (e).

**Figure 4.**
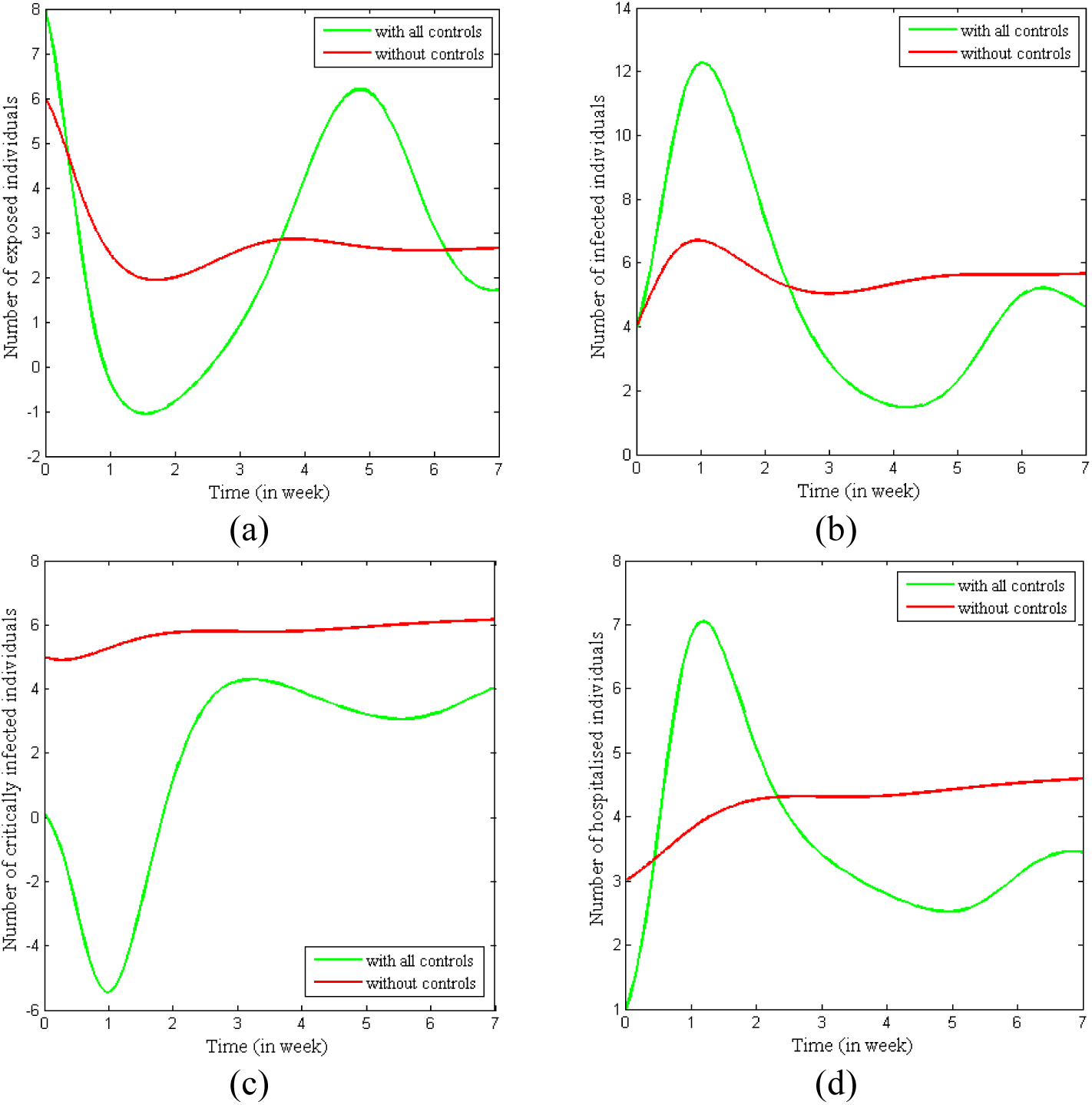
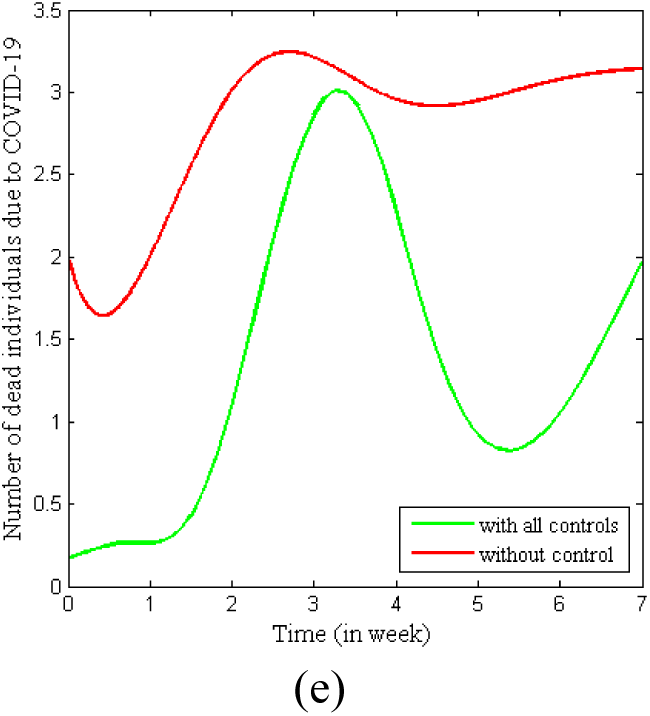
Variation in compartments with and without optimal controls

Variation in intensity of control strategies with time is shown in figure 5. Moreover, the figure also demonstrates which control can be applied at how much intensity to control COVID-19 outbreak in seven weeks. The high fluctuation in *u*_3_ control variable at an initial stage suggests that it is very important to control infected individuals to move at critical stage to reduce mortality due to COVID-19. And, this can be achieved easily if an infected individual gets proper vaccination for this disease. Since effective vaccination is not available for coronavirus, one should take proper care of infected individuals to improve their immunity so that their body becomes capable to fight against the virus and not reaching to a critical stage. Moreover fluctuation in *u*_2_ control variable suggests that it is very important to isolate or quarantine infected individuals to control this pandemic outbreak.

**Figure 5.**
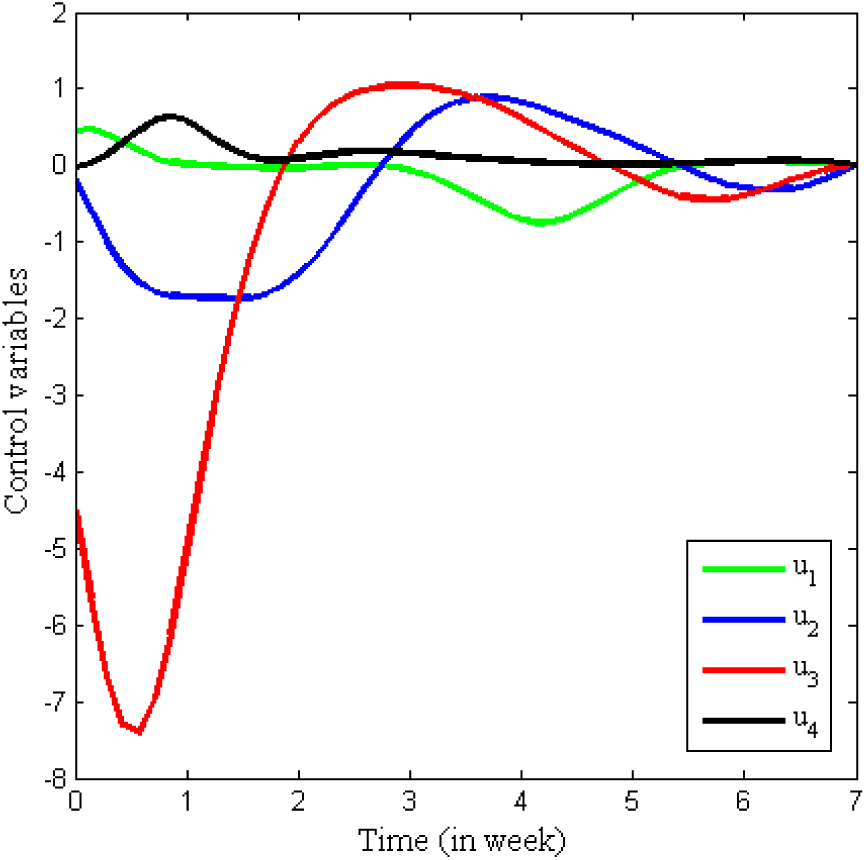
Variation in control variable with time

Separate effect of control variables on each compartment can be observed in figure 6. From the figure, we can interpret that *u*_2_ control variable is highly effective to stabilise this epidemic situation. Figure 6(b) depicts that population class of infected individuals is lowest under the influence of *u*_5_ control variable which suggests that rapid hospitalisation of infected individuals is an effective step to reduce infected cases of COVID-19. Figure 6(c) suggests that to reduce critical cases of COVID-19, first we should make control on infected individuals to become critically infected and we should improve hospitalisation and medical facility for critically infected individuals to save their lives. Figure 6(e) shows that mortality rate due to COVID-19 can be reduced effectively within three weeks of outbreak by applying *u*_1_, *u*_2_ and *u*_3_ control strategies. That means self-quarantine for an exposed individual, isolation of an infected individual and reducing critical cases by taking extra care of infected individuals are effective strategies to control further transmission of COVID-19.

**Figure 6.**
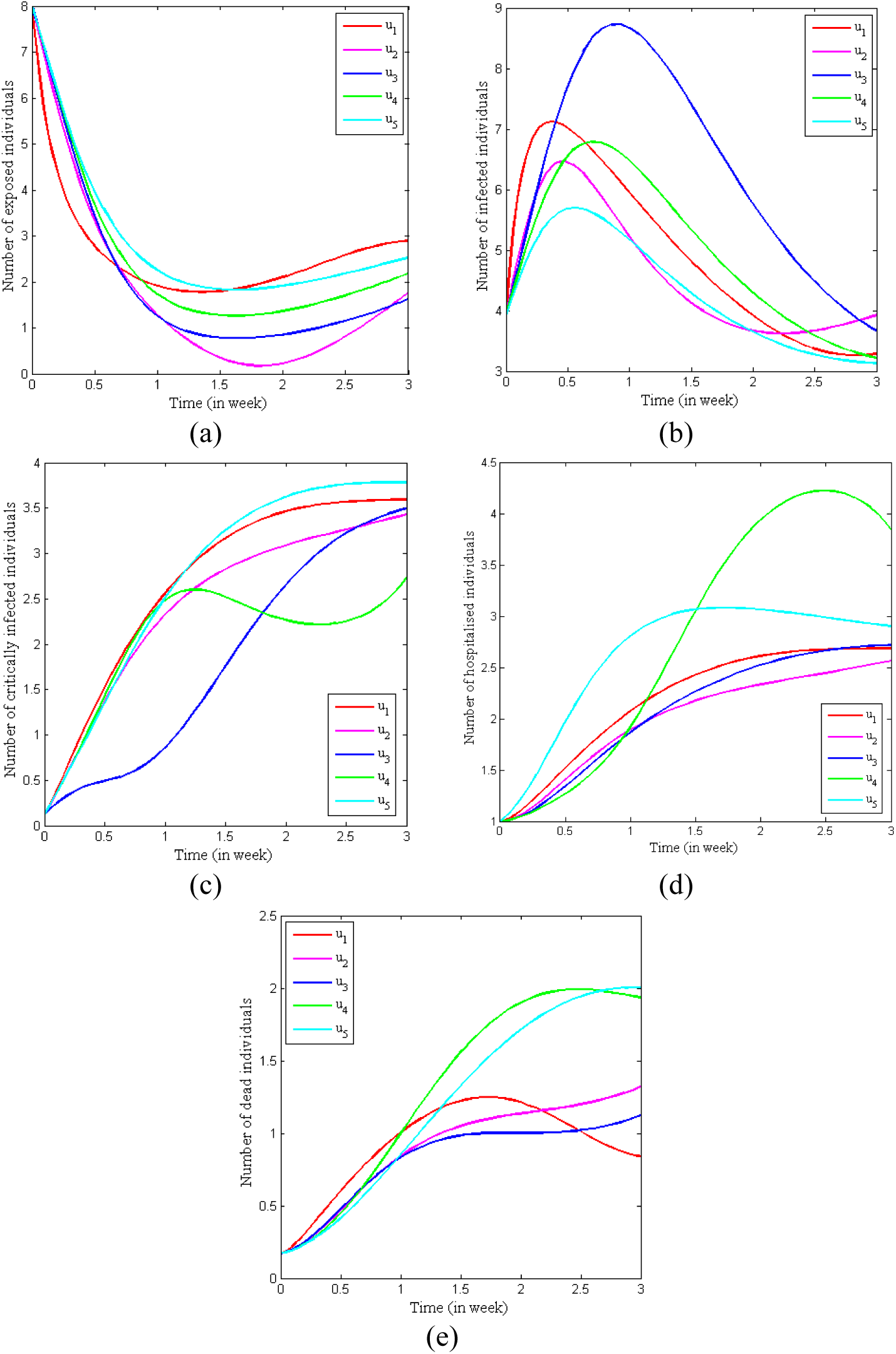
Variation in each compartment under individual effect of control variables

Figure 7 gives change in the objective function under influence of all the controls. Combine effect of all the controls gives fruitful effect on the model.

**Figure 7.**
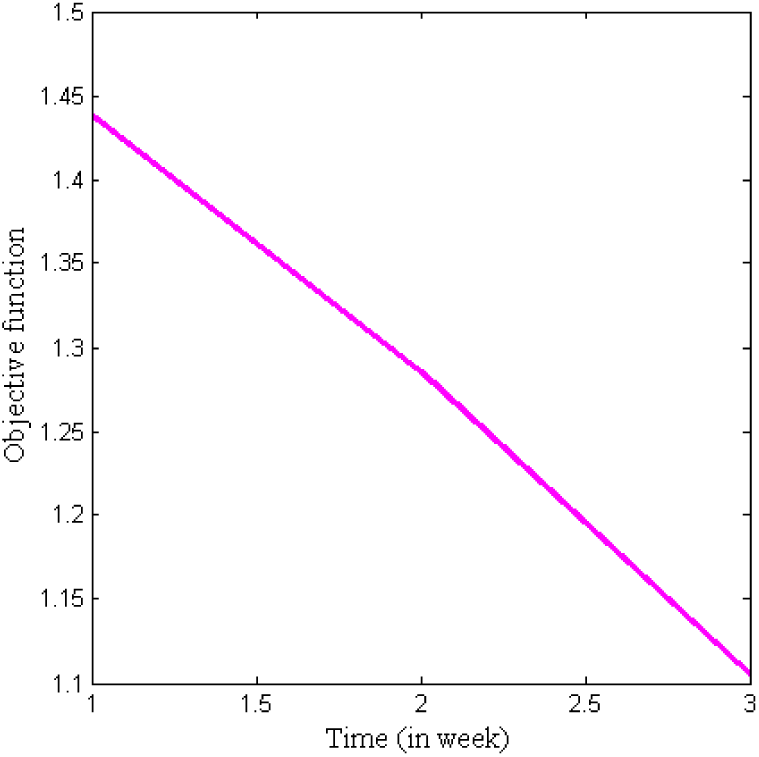
Change in objective function with time

Figure 8 demonstrates the scatter diagram representing chaotic situation created during COVID-19 outbreak. Figure 8(a), 8(b) and 8(c) shows periodic mortality from classes of exposed individuals, infected individuals and critically infected individuals respectively, when under hospitalisation. On comparison of figures 8 (a) and (b), it can be interpreted that mortality ratio in class of infected individuals is higher and much quicker than in the class of exposed individuals. The chaotic figure 8(c) represents a very high mortality rate of critically infected individuals. Hence, in the absence of vaccination for COVID-19, it becomes a challenging situation to cure critically infected individuals.

**Figure 8.**
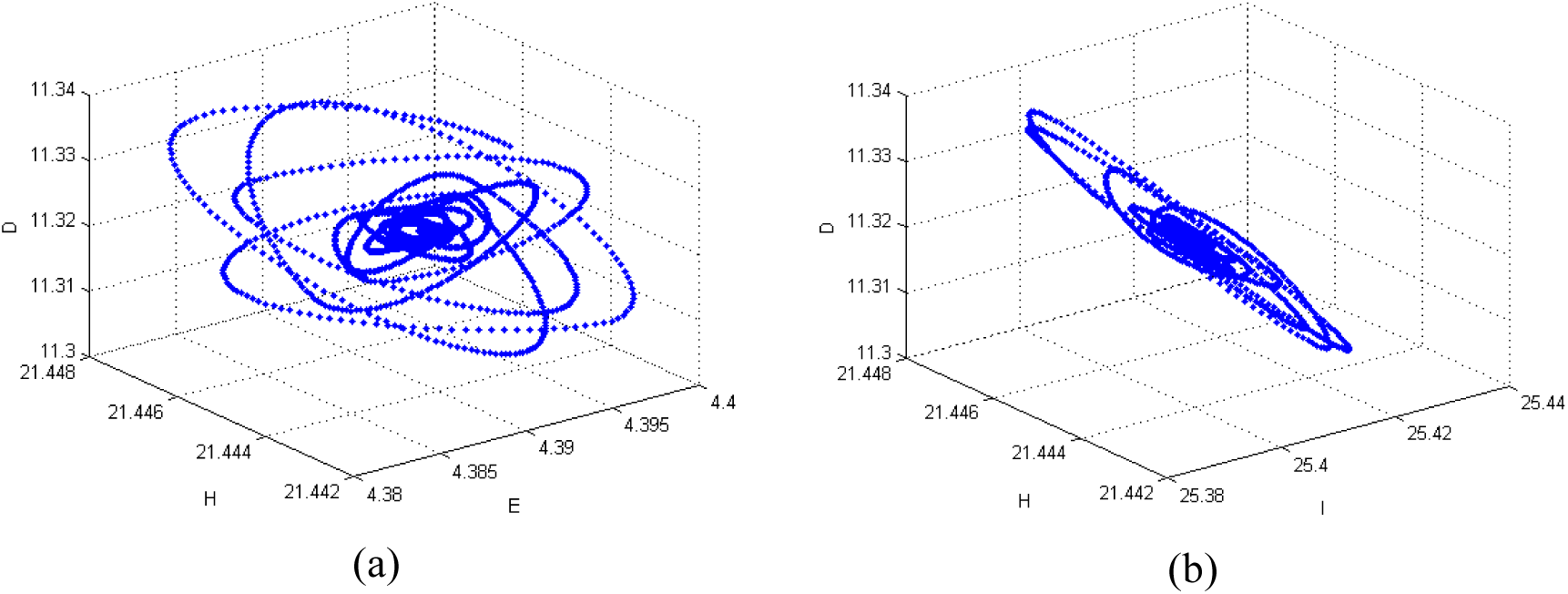
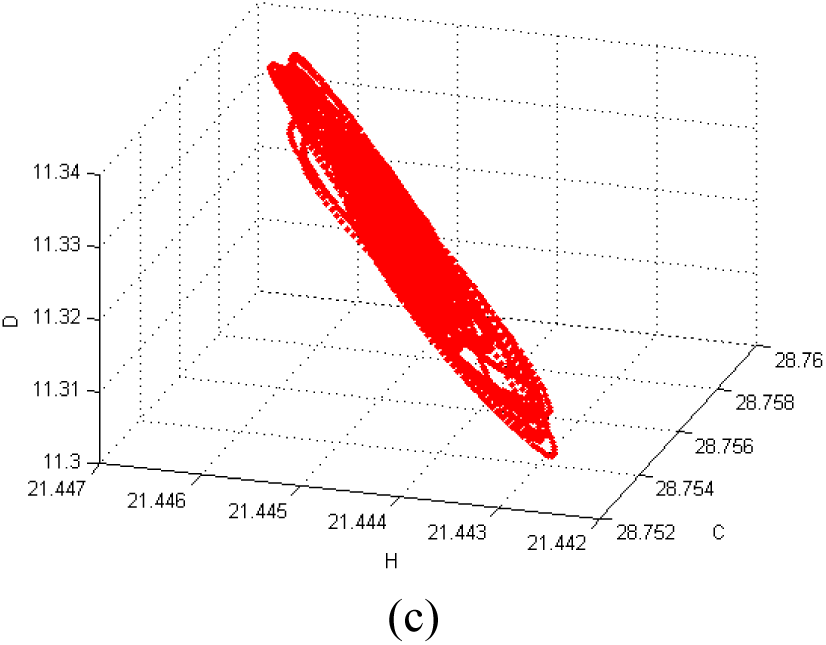
Scatter diagram for COVID-19 outbreak

## 5. Conclusion

In this study, a compartmental model is constructed to examine transmission of COVID-19 in human population class. Moreover, basic reproduction number is formulated to calculate threshold value of the disease. In order to develop strategies to prevent the epidemic of COVID-19, optimal control theory is applied to the model. Further to advance control theory, five control variables are introduced in the model in the form of control strategies. These strategies include self-quarantine of exposed individuals, isolation of infected individuals, taking extra care of infected individuals to reduce critical case of COVID-19, increase hospitalisation facility for infected and critically infected individuals. Distinctive and combined effects of these control variables on all the compartments are observed and examined graphically by simulating the COVID-19 model. Numerical simulation of the model reflects that quarantine and better medical treatment of infected individuals reduce the critically infected cases, which will further reduce the transmission risk and demises.

## Data Availability

Data is included within the manuscript.

## Data Availability

https://www.who.int/emergencies/diseases/novel-coronavirus-2019/situation-reports

## Ackhnowledgement

Second author (AHS) is funded by a Junior Research Fellowship from the Council of Scientific & Industrial Research (file no.-09/070(0061)/2019-EMR-I) and all the authors are thankful to DST-FIST file # MSI-097 for technical support to the Department of Mathematics, Gujarat University. Third author (ENJ) is funded by UGC granted National Fellowship for Other Backward Classes (NFO-2018-19-OBC-GUJ-71790).

## References

[2] Chen, T., Rui, J., Wang, Q., Zhao, Z., Cui, J. A., & Yin, L. (2020). A mathematical model for simulating the transmission of Wuhan novel Coronavirus. bioRxiv.

[3] Chen, Y., Cheng, J., Jiang, Y., & Liu, K. (2020). A time delay dynamic system with external source for the local outbreak of 2019-nCoV. Applicable Analysis, 1–12.

[4] Cheng, Z. J., & Shan, J. (2020). 2019 Novel coronavirus: where we are and what we know. Infection, 1–9.

[5] Diekmann, O., Heesterbeek, J. A. P., & Metz, J. A. (1990). On the definition and the computation of the basic reproduction ratio R0 in models for infectious diseases in heterogeneous populations. Journal of Mathematical Biology, 28 (4), 365–382.

[6] Driessche, P.; Watmough, J.; Reproduction numbers and sub-threshold endemic equilibria for compartmental models of disease transmission, Mathematical Biosciences. 180(1) (2002), 29–48.

[7] Fleming, W., & Lions, P. L. (Eds.). (2012). Stochastic Differential Systems, Stochastic Control Theory and Applications: Proceedings of a Workshop, held at IMA, June 9-19, 1986 (Vol. 10). Springer Science & Business Media.

[8] Khan, M. A., & Atangana, A. (2020). Modeling the dynamics of novel coronavirus (2019-nCov) with fractional derivative. Alexandria Engineering Journal, 1-11.

[9] Khot, W. Y., & Nadkar, M. Y. (2020). The 2019 Novel Coronavirus Outbreak–A Global Threat. J Assoc Physicians India, 68(3), 67–71.

[10] Peng, L., Yang, W., Zhang, D., Zhuge, C., & Hong, L. (2020). Epidemic analysis of COVID-19 in China by dynamical modeling. arXiv preprint 2002.06563.

[11] Riou, J., & Althaus, C. L. (2020). Pattern of early human-to-human transmission of Wuhan 2019 novel coronavirus (2019-nCoV), December 2019 to January 2020. Eurosurveillance, 25(4).

[12] Rothan, H. A., & Byrareddy, S. N. (2020). The epidemiology and pathogenesis of coronavirus disease (COVID-19) outbreak. Journal of Autoimmunity, 102433.

[13] Sahin, A. R., Erdogan, A., Agaoglu, P. M., Dineri, Y., Cakirci, A. Y., Senel, M. E., … & Tasdogan, A. M. (2020). 2019 Novel Coronavirus (COVID-19) Outbreak: A Review of the Current Literature. EJMO, 4(1), 1–7.

[14] Sohrabi, C., Alsafi, Z., O’Neill, N., Khan, M., Kerwan, A., Al-Jabir, A., Iosifidis, C. & Agha, R. (2020). World Health Organization declares global emergency: A review of the 2019 novel coronavirus (COVID-19). International Journal of Surgery, 71–76

[15] Tang, B., Bragazzi, N. L., Li, Q., Tang, S., Xiao, Y., & Wu, J. (2020). An updated estimation of the risk of transmission of the novel coronavirus (2019-nCov). Infectious Disease Modelling, 5, 248–255.

[16] Tang, B., Wang, X., Li, Q., Bragazzi, N. L., Tang, S., Xiao, Y., & Wu, J. (2020). Estimation of the transmission risk of the 2019-nCoV and its implication for public health interventions. Journal of Clinical Medicine, 9(2), 462.

[17] Thevarajan, I., Nguyen, T. H., Koutsakos, M., Druce, J., Caly, L., van de Sandt, C. E., … & Tong, S. Y. (2020). Breadth of concomitant immune responses prior to patient recovery: a case report of non-severe COVID-19. Nature Medicine, 1–3.

[18] World Health Organization. (2020). Coronavirus disease 2019 (COVID-19) : situation report, 51.

[19] Wu, J. T., Leung, K., & Leung, G. M. (2020). Nowcasting and forecasting the potential domestic and international spread of the 2019-nCoV outbreak originating in Wuhan, China: a modelling study. The Lancet, 395(10225), 689–697.

[20] Yang, C., & Wang, J. (2020). A mathematical model for the novel coronavirus epidemic in Wuhan, China. Mathematical Biosciences and Engineering, 17(3), 2708–2724.

[21] Zhao, Z., Zhu, Y. Z., Xu, J. W., Hu, Q. Q., Lei, Z., Rui, J., Liu, X., Wang, Y., Luo, L., Yu, S.S. & Li, J. (2020). A mathematical model for estimating the age-specific transmissibility of a novel coronavirus. medRxiv,.

[22] Zhong, L., Mu, L., Li, J., Wang, J., Yin, Z., & Liu, D. (2020). Early Prediction of the 2019 Novel Coronavirus Outbreak in the Mainland China based on Simple Mathematical Model. IEEE Access, 51761-51769.

[23] https://www.who.int/emergencies/diseases/novel-coronavirus-2019/situation-reports

